# Effects of Soil Moisture and Soil Temperature on Coccidioidomycosis

**DOI:** 10.1101/2025.06.30.25330594

**Authors:** Qianqian Li, Beichen Zhang, Runqiu Wang, Haiyue Li, Yue Zhan, Daniel Tong, Jesse E. Bell

## Abstract

**Key Points:**

- Multi-year soil wet–dry and cool–hot cycles along with concurrent dry, dusty conditions, are linked to higher Valley fever incidence across exposure seasons
- Higher incidence is linked to prior wetter winters and monsoons, and hotter, drier falls and springs, with winter and spring soil conditions most influential
- Results call for adding soil indicators with up to 3-year lead times into early-warning systems to enhance public health actions in endemic regions

Coccidioidomycosis (Valley fever, VF) is a climate-sensitive infectious disease caused by inhaling soil-dwelling fungus *Coccidioides*, mostly reported in southwestern USA. Although soil moisture (SM) and soil temperature (ST) are known to shape the fungal lifecycle, their effects on coccidioidomycosis remain understudied. Most prior studies have relied on their proxies— precipitation and air temperature—that might not accurately capture soil hydrothermal dynamics. We conducted multivariable negative binomial regressions to estimate seasonal associations between incidence and climate drivers—including SM, ST, and wind speed from the North American Land Data Assimilation phase 2 (NLDAS-2), and PM_10_-based dusty-day counts—in Arizona’s hyperendemic counties (Maricopa, Pima, and Pinal) from 2000 to 2022. We found higher incidence in areas with hotter, drier soils and more seasonal dusty days. Multi-year soil hydrothermal cycles—alternating wet–dry and cool–hot periods along with concurrent dry, dusty conditions—significantly influenced incidence. Notably, no antecedent dry–cool seasons were linked to increased incidence, indicating moisture or heat as prerequisites for fungal growth or dispersal. SM showed more consistent and widespread effects than ST across seasons and lags, with winter and spring soils most influential. Higher incidence followed wetter winters and monsoons, and dry, hot springs and falls. Our models using NLDAS-2 SM and ST data showed robust performance and generalizability across exposure seasons. Our results support adding multi-year soil indicators—with up to three-year lead times—into early-warning systems to enhance VF forecasting and better prepare endemic regions for the challenges of a warming, drying, and increasingly variable climate.

**Plain Language Summary:** Coccidioidomycosis is a fungal disease caused by inhaling *Coccidioides* spores from soils, most commonly in southwestern USA. Soil moisture and temperature are known to influence the fungal lifecycle, but their roles remain poorly understood because most studies used their proxies—precipitation and air temperature—that might not accurately reflect soil water-temperature patterns. We examined how current wind, dusty days, and both current and past soil moisture and soil temperature influenced incidence. We found more cases in Arizona regions with drier, hotter soils and more seasonal dusty days. Importantly, alternating wet–dry and cool– hot soil cycles over the past 0–3 years significantly influenced disease occurrence. No past dry– cool conditions were linked to higher incidence, suggesting either moisture or heat is needed for fungal proliferation or dispersal. Soil moisture had more consistent and extensive seasonal effects than soil temperature. Higher incidence was linked to wetter soils in prior winters and monsoons, and hotter, drier soils in falls and springs, with winters and springs most influential. Our models using reanalysis-based soil data showed robust performance across seasons. Our findings support adding soil data with up to three-year lead times into early-warning systems to improve public health actions in endemic regions.

## 1. Introduction

Coccidioidomycosis (Valley fever, VF), is an infectious fungal disease caused by inhaling airborne arthroconidia (i.e., spores) of *Coccidioides immitis and C. posadasii*. These fungi thrive in alkaline, arid to semiarid thermic soils throughout the southwestern USA, southcentral Washington state, northern Mexico, and parts of Central and South America (Ashraf et al., 2020; Crum, 2022; del Rocío Reyes-Montes et al., 2016; Fisher et al., 2007; Kollath et al., 2019; Maddy, 1958). *C. immitis* prevails in California and Washington state, while *C. posadasii* dominates the remaining endemic regions (Ashraf et al., 2020). *Coccidioides* displays a dimorphic lifecycle, existing in a saprobic (soil) and a parasitic (host) phase (Nguyen et al., 2013). In soils, it grows as filamentous mycelia, often isolated at depths of 2–20 cm (Fisher et al., 2007), which fragment into arthroconidia under sufficiently dry conditions. These arthroconidia are easily aerosolized by wind erosion or other soil disturbances and delivered through airborne dust, or—in rare cases—through direct contact between soils and open wounds (Litvintseva et al., 2015). After being inhaled by a susceptible host, the fungus will enter its parasitic phase, developing spherules in the lungs that release numerous small endospores, thus sustaining the infection cycle. About 40% of infections become symptomatic within 1–3 weeks post-exposure, with 5–10% developing severe or chronic forms and 0.5–2% progressing to disseminated disease (Crum, 2022; Donovan et al., 2025; Toda et al., 2024). Although Valley fever is not transmitted between individuals, the absence of an effective vaccine or definitive treatment and the challenges in diagnosis and management make it an essential public health concern in endemic areas (Crum, 2022; Donovan et al., 2024).

Over recent decades, VF incidence has increased significantly and is expected to rise further in endemic regions, coinciding with the expanding geographic range of *Coccidioides* endemicity (Ashraf et al., 2020; CDC Wonder, 2021, 2024; Centers for Disease Control and Prevention, 2024; Donovan et al., 2024; Gorris et al., 2019; Head et al., 2022; Hepler et al., 2025; Howard et al., 2024; S. L. Williams & Chiller, 2022). From 1998 to 2023, annual reported cases in the U.S. increased more than ninefold, with Arizona’s cases surging from 1,474 to 10,990 and accounting for 61% of nearly 300,000 total nationwide cases—almost double California’s 36% (Arizona Department of Health Services, 2024b; California Department of Public Health, 2024; Centers for Disease Control and Prevention, 2024). About 95% of Arizona’s cases were reported in the hyperendemic southcentral region (Figure 1a), which includes Maricopa, Pima, and Pinal Counties and spans the Phoenix and Tucson metropolitan areas. In this region, Valley fever has become a leading cause of community-acquired pneumonia, resulting in severe morbidity and substantial economic burdens (Arizona Department of Health Services, 2024b; Crum, 2022; Grizzle et al., 2021; Kim et al., 2009; Valdivia et al., 2006). Evidence increasingly suggests that changing climate conditions contribute significantly to this escalating public health threat (Comrie, 2005; Donovan et al., 2024; Gorris et al., 2018, 2019; Head et al., 2022; Howard et al., 2024; Kolivras & Comrie, 2003; Lee et al., 2025; Roach et al., 2017; E. Weaver & Kolivras, 2018).

**Figure 1.**
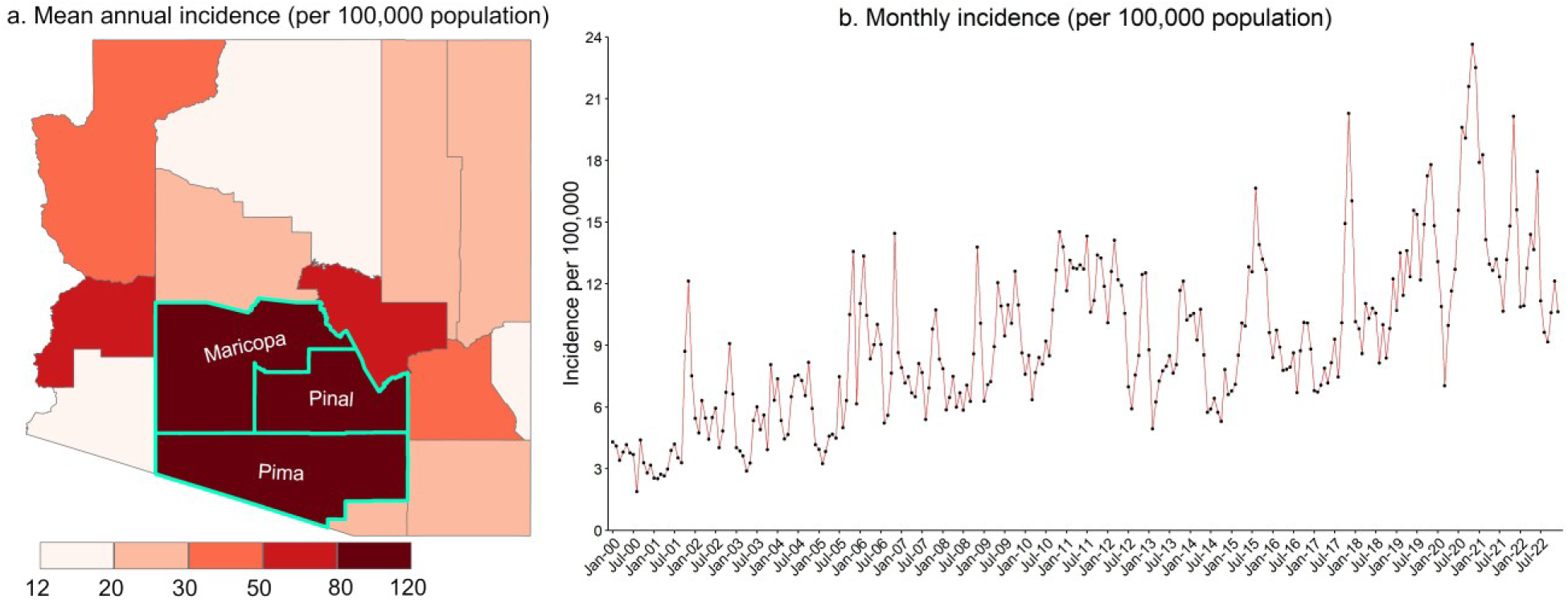
Spatial and monthly trends in coccidioidomycosis incidence, 2000–2022. (a) Mean annual coccidioidomycosis incidence by county across Arizona, (b) Monthly coccidioidomycosis incidence in the study region, which includes Maricopa, Pima, and Pinal counties (outlined in green in map (a)) and was used for all analyses. Note that incidence data were adjusted to account for a 1-month reporting delay and for artificial case inflation between June 2009 and November 2012 by halving case counts during that period.

Arizona reports the highest VF case numbers, with a noticeably increasing trend in recent decades, which is likely driven by its unique and evolving climate conditions. Situated in the hottest and driest region of the United States, Arizona—particularly its southcentral region—is characterized by expansive, wind-erodible, and human-disturbed arid and semiarid drylands and deserts (Achakulwisut et al., 2019; A. P. Williams et al., 2022). In recent decades, the Southwest has experienced rising temperatures with more frequent hot days, declining overall precipitation with more heavy rainfall events, increasingly severe and prolonged droughts, and dustier conditions (Pinson et al., 2023; Su et al., 2023; Tong et al., 2017; White et al., 2023; A. P. Williams et al., 2022). A megadrought beginning in 2000 has led to the driest multidecadal period in Arizona for at least 1,200 years, as evidenced by severe soil moisture deficits exacerbated by anthropogenic climate change (A. P. Williams et al., 2020, 2022). Climate projections indicate continued warming, reduced water availability, increased drought severity, and elevated dust emissions (Achakulwisut et al., 2019; Cook et al., 2015, 2021; Pinson et al., 2023, 2023; Strzepek et al., 2010; Su et al., 2023; Tong et al., 2017; White et al., 2023; A. P. Williams et al., 2022). These climatic changes may have contributed to the increasing VF incidence and expanding endemic areas and are expected to continue driving VF dynamics in the future (Gorris et al., 2018, 2019; Head et al., 2022; Hepler et al., 2025; Howard et al., 2024; Kumar et al., 2024; Lee et al., 2025; A. K. Weaver et al., 2025).

The distribution and abundance of *Coccidioides* spp. in the soil and atmosphere are thought to be influenced by climate conditions through their effects on the fungal lifecycle —including growth, sporulation, and aerosolization. However, direct environmental detection of *Coccidioides* remains challenging, limiting the availability of long-term time series fungal data for robust climate impact assessments. Consequently, prior studies have relied on VF incidence data, linking it to climate conditions under the assumption that factors influencing fungal growth and dispersion also affect disease occurrence in endemic regions (Comrie, 2005; Gorris et al., 2018; Head et al., 2022; E. Weaver & Kolivras, 2018). The prevailing mechanistic theory, the “grow and blow” hypothesis, suggests that *Coccidioides* require sufficient soil moisture provided by seasonal precipitation and favorable temperatures to support hyphal growth and proliferation, while subsequent hot, dry conditions promote hyphal desiccation and fragmentation into infectious arthroconidia, which can become airborne and dispersed through dust when soil is disturbed by wind or other mechanisms, leading to potential inhalation (Comrie, 2005; Comrie & Glueck, 2007; Fisher et al., 2007; Tamerius & Comrie, 2011; E. Weaver & Kolivras, 2018).

Another widely accepted “soil sterilization” hypothesis posits that *Coccidioides*, as a filamentous organism, can survive extreme hot and dry conditions by extending its hyphae into deeper soil layers—typically characterized by greater moisture and lower temperature than the topsoil—to evade non-filamentous microbial competitors; subsequently, when favorable conditions return, it thrives in the topsoil with few competing microorganisms (Fisher et al., 2007; Maddy, 1958, 1965). Previous studies have examined the effects of concurrent or antecedent precipitation and air temperature on the seasonality and interannual variability of VF incidence (Comrie, 2005; Gorris et al., 2018; Head et al., 2022; Kolivras & Comrie, 2003; Tamerius & Comrie, 2011). In Arizona, precipitation during the typically arid foresummer (April–June) has been associated with increased VF incidence across all seasons 1.5–2 years later (Comrie, 2005), whereas concurrent precipitation was negatively correlated with incidence, likely due to suppressed spore dispersal (Tamerius & Comrie, 2011). Kolivras and Comrie (2003) highlighted the importance of winter climatic conditions, noting that winter precipitation and temperature were particularly influential, with greater effects on annual incidence than other seasons. Specifically, they observed that the winter months (January–March) occurring 2 years before a high-incidence January experienced below-average temperatures and above-average precipitation. In California, Head et al. (2022) found that VF incidence peaked at the end of hot and dry seasons, particularly following anomalously wet winters. Their findings also indicated that wetter winter and spring months followed by hotter, drier summer months were associated with higher fall incidence. A broader investigation by Gorris et al. (2018) found that higher VF incidence across the southwestern United States was linked to hotter, drier, and dustier conditions, and in California’s San Joaquin Valley, incidence in the fall was increased in years preceded by cool, wet spring seasons.

Despite soil moisture (SM) and soil temperature (ST) are known to influence *Coccidioides* lifecycle, most prior studies have relied on precipitation and air temperature (AT) as proxies to evaluate climatic effects on VF incidence. However, these proxies may not adequately reflect soil hydrothermal conditions where the fungus grows, potentially introducing uncertainties into climate-based VF risk assessments. Although high-quality in situ observations of SM and ST with sufficient temporal and spatial coverage for reliably analyzing their effects on VF incidence remain unavailable, model-based datasets—such as the Noah simulations from Phase 2 of the North American Land Data Assimilation System (NLDAS-2)—have become increasingly accessible over the past decade, providing continuous, multi-year data across broad geographic regions. These model-based SM products offer a more reliable indicator of root-zone water availability than precipitation alone, as they are simulated by incorporating a range of interacting drivers that influence soil water balance—including precipitation, evapotranspiration, infiltration, runoff, solar radiation, temperature, soil texture, and vegetation type (Xia et al., 2015b)—all of which collectively determine soil water retention critical for hyphal growth and mycelial proliferation. As a comprehensive integrator of these factors, the Noah-simulated SM in NLDAS-2 serves as a superior indicator of drought conditions than precipitation alone and more effectively captures the soil wet–dry cycles underpinning the widely accepted “grow and blow” hypothesis. Similarly, the Noah-simulated ST provides a more reliable metric of root-zone thermal conditions than AT alone and may be more directly relevant to, and influential on, the lifecycle of *Coccidioides*. Although ST is influenced by AT, substantial differences between the two can arise depending on soil depth and various environmental influences (Fisher et al., 2007). Field measurements have shown that even under high AT and potentially lethal soil surface temperatures, ST below the surface (e.g., at 20 cm depth) can remain within the favorable growth range (20–40℃) for *Coccidioides* (Fisher et al., 2007). In addition to AT, the Noah-simulated ST is influenced by a suite of interacting factors, including solar radiation, soil heat flux, soil thermal conductivity, soil heat capacity, soil texture, vegetation type, evapotranspiration, and soil moisture (Xia et al., 2015b). As a result, it more accurately characterizes soil thermal dynamics than AT alone, thereby more effectively capturing the hot–cool cycles relevant to both the “soil sterilization” and “grow and blow” hypotheses. Therefore, incorporating model-based SM and ST data is essential for advancing our understanding of coccidioidomycosis ecology and for improving predictions of disease incidence in response to climatic variability.

Earlier studies often relied on AT as a proxy for ST, and on precipitation or other surrogates such as the Normalized Difference Vegetation Index (NDVI) as substitutes for SM (Stacy et al., 2012). The effects of SM and ST on VF incidence remain underexplored. Only a few recent efforts (Coopersmith et al., 2017; Gorris et al., 2018) have begun to incorporate SM data; however, these studies relied primarily on correlation or univariate analyses, thereby neglecting potential confounding effects from concurrent and lagged SM or ST time series, nor from other covariates such as wind speed or PM_10_. Such reliance on univariable rather than multivariable frameworks limits the ability to isolate the individual contributions of an exposure variable from the combined effects of its covariates, potentially oversimplifying the complex relationships that influence VF incidence. Moreover, no study to date has quantitatively investigated the relationship between concurrent and lagged ST and VF incidence. In this study, we aim to address these critical knowledge gaps by conducting the first comprehensive multivariable modeling analysis to investigate the concurrent and lagged effects of SM and ST on incidence. Additionally, our analytical framework incorporates indicators of soil disturbance (wind speed) and airborne dust (PM_10_, inhalable particles ≤10 μm in diameter, used as a proxy for airborne arthroconidia concentration), as suggested by the dominant “grow and blow” mechanistic hypothesis. Prior studies have shown significant associations between these two factors and VF incidence (Comrie, 2005; Jin et al., 2025; Kollath et al., 2022; E. Weaver & Kolivras, 2018), indicating that they may modify how soil moisture and soil temperature influence disease incidence. By leveraging publicly available model-based SM and ST data to evaluate their concurrent and lagged seasonal effects on VF incidence, this study seeks to enhance our understanding of prevailing hypotheses by comparing our findings with prior studies that relied on SM or proxies such as precipitation, NDVI, or AT, thereby contributing to improved disease prediction and public health mitigation in vulnerable regions.

## 2. Methods

### 2.1. Coccidioidomycosis Data

This longitudinal ecological study used monthly, county-level coccidioidomycosis case counts— by county of residence—in Arizona from January 2000 through December 2022, obtained from the Arizona Department of Health Services (ADHS; Table 1). Cases are recorded by the month and year of their report date (i.e., the date of laboratory reporting to ADHS); since 1997, all healthcare providers and laboratories in Arizona have been mandated to report confirmed coccidioidomycosis cases to ADHS within 5 working days of laboratory confirmation (Arizona Department of Health Services, 2018). A 1-month offset was applied to the case data to account for a sequence of delays between exposure and report date, including the incubation period from spore exposure to symptom onset, the time required for diagnosis, and subsequent laboratory testing and reporting lags (Comrie & Glueck, 2007). The reported case counts between June 2009 and November 2012 were divided by two to adjust for the near doubling in cases attributed to changes in laboratory reporting and testing practices (Arizona Department of Health Services, 2018, 2024a; Comrie, 2021). County-level annual mid-year population estimates for 2000–2022 were obtained from the Surveillance, Epidemiology, and End Results (SEER) Program of the National Cancer Institute (SEER, 2025). These annual population data were linearly interpolated to generate monthly population estimates, which were subsequently used to calculate monthly county-level coccidioidomycosis incidence rates per 100,000 population. Our analyses focused on the hyperendemic region including Maricopa, Pima, and Pinal counties, which accounted for about 95% of Arizona’s reported cases (158,707 out of 166,767 statewide) during the 23-year study period (Figure 1a). Monthly incidence in the hyperendemic region was plotted over the study period, with adjustments for inflated case counts reported from 2009–2012 (Figure 1b).

**Table 1.** List of Data Used in This Study.

| Category | Variables | Spatial/Temporal resolution | Period | Source |
| --- | --- | --- | --- | --- |
| Exposures | Soil moisture (0–10 cm)<br>(kg m<sup>-2</sup>) | 0.125°×0.125°/Monthly | 1997–2022 | North American Land Data Assimilation System (NLDAS), Noah land surface model (LSM), Version 2.0 |
|  | Soil temperature (0–10 cm) (K) | 0.125°×0.125°/Monthly | 1997–2022 | NLDAS, Noah LSM, Version 2.0 |
|  | Wind speed at 10 m above ground (m s<sup>-1</sup>) | 0.125°×0.125°/Monthly | 1997–2022 | NLDAS, Primary forcing data, Version 2.0 |
|  | Daily mean PM<sub>10</sub> concentration (µg m<sup>-3</sup>) | Monitoring site/Daily | 1997–2022 | U.S. Environmental Protection Agency, Outdoor air quality data |
| Outcome | Valley fever case count | County/Monthly | 2000–2022 | Arizona Department of Health Services |

### 2.2. Climate Data

Prior studies have shown that climate conditions may influence coccidioidomycosis incidence with delays of up to three years (Comrie, 2005; Head et al., 2022). Accordingly, we collected climate data from 1997 to 2022 (Table 1), and all analyses were conducted using climate data over this period.

Monthly gridded estimates of soil moisture, soil temperature, and wind speed at a 0.125° spatial resolution were obtained from NLDAS-2 (NLDAS project, 2022a, 2022b; Xia et al., 2012). Specifically, the SM and ST data were simulated from the Noah land surface model (LSM) for NLDAS-2, which assimilated observations of precipitation, air temperature, surface pressure, and solar radiation, along with various soil and vegetation parameters derived from observations or reanalysis products (Xia et al., 2012, 2015b, 2015a). These model-simulated SM and ST are generated through an interacting and iterative modeling process in which each variable dynamically influenced and fed back upon the other, thereby coupling the water and energy cycles through a series of complex linear and nonlinear physical processes (Xia et al., 2015b).

The resulting SM and ST products reflect the interactions among a range of hydrological and thermal drivers—such as precipitation, air temperature, evapotranspiration, runoff, infiltration, radiation, and soil and vegetation properties—that jointly govern soil hydrothermal dynamics (Xia et al., 2015b). The performance of these products has been comprehensively evaluated across multiple soil depths (0–10 cm, 10–40 cm, 40–100 cm) and temporal scales (daily, monthly, annual) through comparisons with in situ observations (Xia et al., 2013, 2014). Xia et al. (2014) demonstrated that the Noah model can effectively capture broad patterns of observed SM variability, including seasonal cycles, interannual fluctuations, and wet/dry events across depths and timescales. This strong performance makes Noah-simulated SM particularly well-suited for this study, given the central role of soil wet–dry cycles in the widely accepted “grow and blow” hypothesis. Similarly, Xia et al. (2013) showed that Noah-simulated ST aligns well with observed values, particularly in the upper soil layers, across multiple timescales. In this study, we focus on SM and ST in the surface soil (0–10 cm) layer, because this topsoil layer is most directly involved in—and sensitive to—soil disturbances that facilitate spore dispersal. For wind speed, we used the 10-m above-ground primary forcing data of NLDAS-2, which are derived from the North American Regional Reanalysis (NARR) (Xia et al., 2012). This dataset includes two near-surface (10-m) wind components: zonal (west–east) and meridional (south– north). The total 10-meter wind speed was calculated as the square root of the sum of the squares of the zonal and meridional components. To match the spatiotemporal resolution of the county-level coccidioidomycosis case counts, all gridded climate variables (SM, ST, and wind speed) were spatially averaged by county and linked to the corresponding monthly case data.

For airborne dust, we used daily mean concentrations of PM_10_ measured at outdoor air monitoring sites, obtained from the U.S. Environmental Protection Agency (EPA)’s Outdoor Air Quality Data portal (EPA, 2024). We defined a “dusty day” for a county as any day on which the daily mean PM_10_ concentration exceeded 45 µg m^-3^ at one or more monitoring sites within that county. Conversely, a “non-dusty day” was defined as any day when all monitoring sites within the county recorded PM_10_ concentrations at or below 45 µg m⁻³. This threshold is consistent with the recommended World Health Organization (WHO)’s global air quality guideline (AQG) level (World Health Organization, 2021). We then calculated the monthly total count of dusty days for each county and linked it to the case data for analysis.

### 2.3. Analyses

#### 2.3.1. Seasonal Grouping

Before estimating the effects of concurrent and lagged soil moisture and soil temperature on coccidioidomycosis incidence, we aggregated the monthly data into seasonal groups. As originally described by Comrie (2005) and later reiterated by Weaver and Kolivras (2018), this seasonal analysis offers several benefits: (a) it effectively partitions the year into alternating wet and dry periods, (b) it aids in the detection of critical recurring periods throughout the year, (c) it simplifies the analysis by reducing the complexities introduced by monthly variability, (d) it helps mitigate potential bias from exposure date misclassification, and (e) it enables a clearer, more straightforward evaluation and interpretation of seasonal lag relationships. Seasonal groupings were defined by centering on the periods of maxima and minima in mean monthly coccidioidomycosis incidence and climate drivers, as shown in Figure 2. The data were then categorized into four seasons: winter (January–March), spring (April–June), monsoon (July– September), and fall (October–December). This categorization was chosen because these seasonal groups captured the most significant peaks and troughs in the data—specifically, one maximum and two minima in both incidence and dusty days, as well as one maximum for soil moisture and one minimum for wind speed. Using consistent seasonal groupings across the data facilitates clearer, more straightforward interpretation and enables more intuitive analysis of seasonal climate effects, while also making comparisons with other studies easier. Each season was analyzed individually because climate conditions can vary substantially across seasons (Comrie, 2005; Kolivras & Comrie, 2003), potentially leading to differing effects on incidence by season.

**Figure 2.**
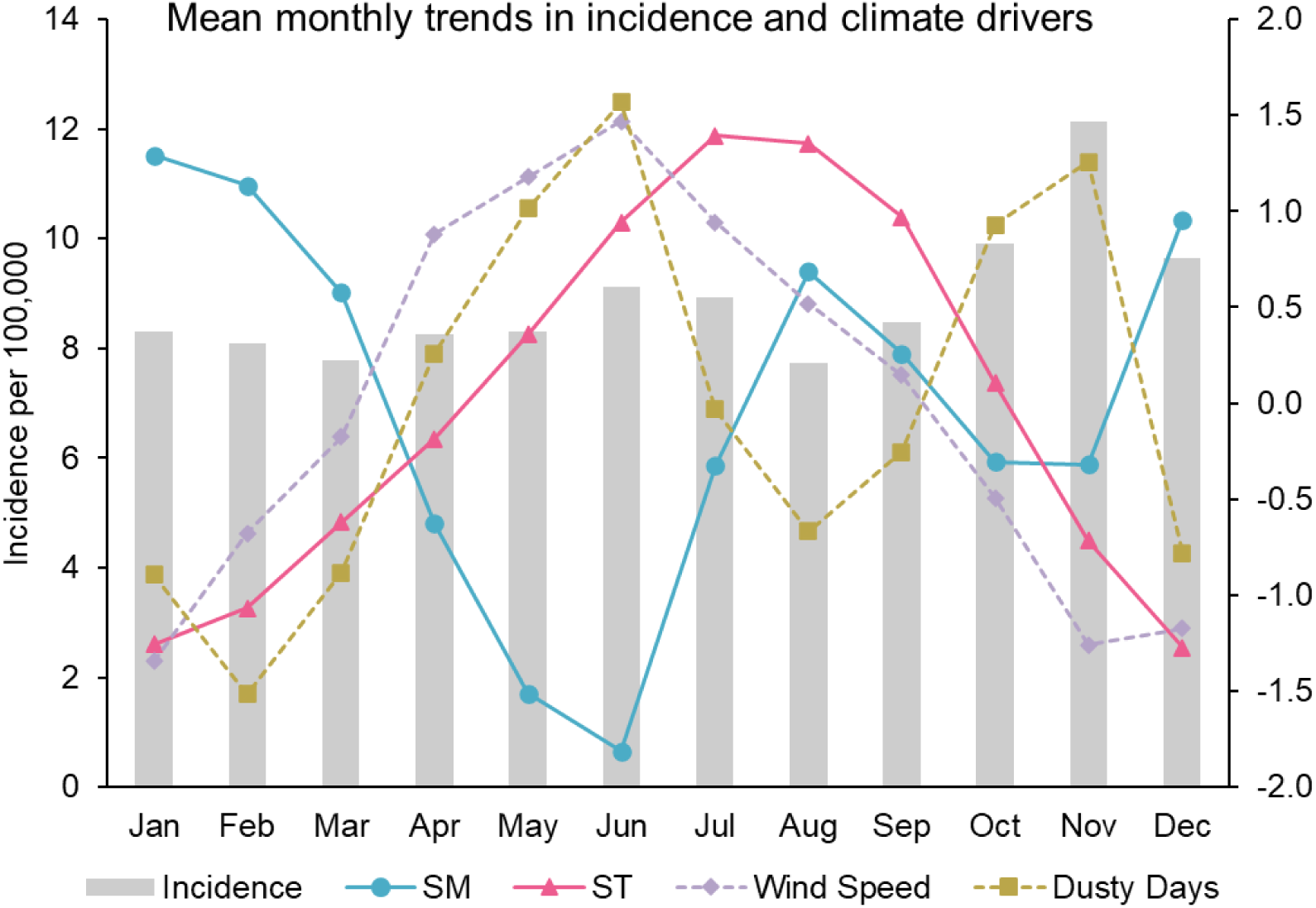
Mean monthly coccidioidomycosis incidence and climate drivers in the study region. Mean monthly incidence (2000–2022) is shown alongside standardized monthly climate variables (1997–2022): soil moisture (SM; 0–10 cm), soil temperature (ST; 0–10 cm), 10-m wind speed, and dusty-day counts. Incidence data were adjusted for 1-month reporting delays and case inflation (Jun 2009–Nov 2012).

#### 2.3.2. Statistical Analysis

Separate multivariable negative binomial regressions (MNBRs) were conducted for each season to estimate the effects of both concurrent (wind speed and number of dusty days) and concurrent and lagged soil moisture and soil temperature on coccidioidomycosis incidence. Negative binomial regression was used to accommodate overdispersion in disease count data. In the multivariable analysis, adjusted incidence rate ratios (aIRRs) and their 95% confidence intervals (CIs) were calculated to quantify the effect of predictors while controlling for other covariates.

Prior to regression analysis, missing covariate data were imputed using an autoregressive integrated moving average (ARIMA)-based time series approach (Hyndman & Athanasopoulos, 2018). Notably, only one covariate (“number of dusty days”) had missing values (∼2.9% of its observations); all other variables were complete. After imputation, bivariate relationships among covariates were assessed using Spearman correlations. These correlation analyses indicated potential multicollinearity among explanatory variables. We combined the three adjacent hyperendemic counties for our analysis because they exhibited similar incidence and predictor patterns as a spatial cluster (Figures 1a and 3), and exploratory analyses showed no significant between-county variability in exposure–outcome relationships. This approach increased our sample size from 23 to 69, enabling multivariable analysis with 28 explanatory variables and substantially improving statistical power and estimate reliability. Initial seasonal MNBR models that included all predictors confirmed substantial multicollinearity, as indicated by variance inflation factor (VIF) values exceeding 10—a common threshold signifying problematic multicollinearity (Vittinghoff et al., 2006). Such high multicollinearity can inflate standard errors and lead to unstable coefficient estimates, underscoring the need for careful variable selection.

**Figure 3.**
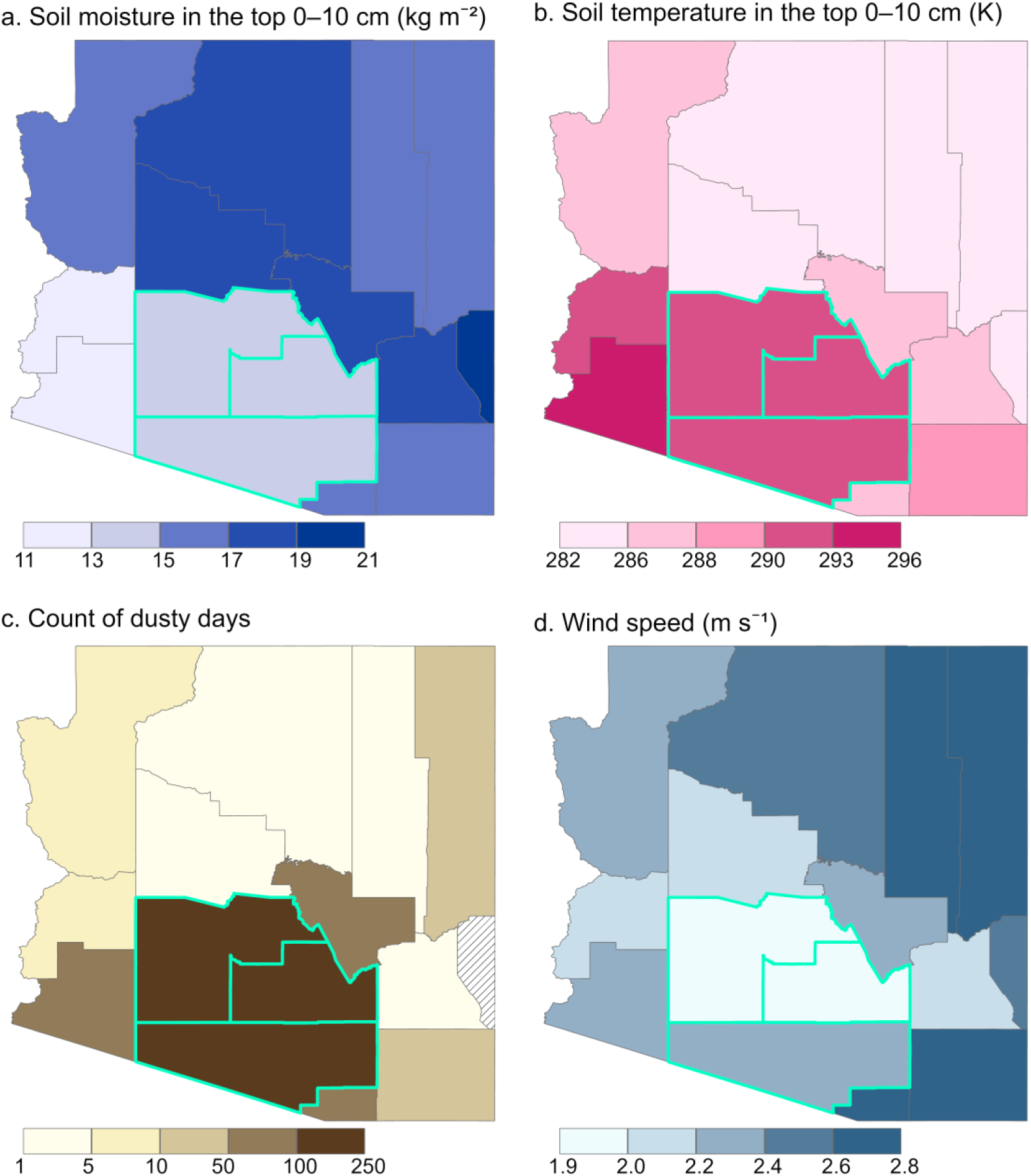
Mean annual spatial patterns of climate drivers, 1997–2022. (a) Average soil moisture in the surface soil layer (topsoil, 0–10 cm), (b) average soil temperature in the surface soil layer (topsoil, 0–10 cm), (c) annual count of dusty days, (d) average wind speed at 10 m above ground. In (c), a “dusty day” for a county was defined as any day on which the daily mean PM_10_ concentration exceeded 45 µg m^-3^ at one or more monitoring sites within that county. Note that counties with diagonal hatching indicate areas with no available data. The study region is outlined in green.

To address multicollinearity, a two-stage variable selection strategy was implemented. First, candidate predictors were screened individually via univariate negative binomial regressions (UNBRs); variables not significantly associated with incidence (2-sided *p-*value ≥ 0.1) were considered for exclusion (Bailey & Gribskov, 1998; Rovetta et al., 2025). However, few variables were eliminated at this stage. Consequently, a second-stage selection was conducted using a restricted bidirectional stepwise procedure—referred to as the Keeping-Exposure Stepwise Selection (KESS) approach—which forced the primary exposure variable to remain in the model at each step, while allowing all other covariates underwent stepwise entry and removal based on the minimum Akaike Information Criterion (AIC) (Harrell, 2015). This approach yielded final models in which no covariate had a VIF > 10, and the resulting aIRRs (with 95% CIs) for the primary exposure were reported.

In addition to the inferential models assessing climate effects, we developed season-specific predictive models using an unrestricted bidirectional stepwise selection procedure applied to each MNBR, selecting predictors solely based on the minimum AIC without forcing any variables to remain in the model. Predictive performance was then assessed using leave-one-out cross-validation (LOOCV), wherein the model was iteratively trained on all observations except one and tested on the held-out case. Standardized regression coefficients (β) were calculated for all climate exposures estimated by the KESS-selected inferential MNBR models, as well as for variables retained in the seasonal predictive models, using z-score–standardized predictors.

Within each seasonal predictive model, the absolute value of β enables direct comparison of the relative influence of predictors measured on different scales. Larger |β| values indicate greater influence on incidence prediction. Notably, standardizing variables does not alter their statistical significance or overall model performance—the *p*-values, coefficient direction (positive/negative), and fit metrics such as AIC, coefficient of determination (R^2^), adjusted (adj.) R^2^, Nagelkerke (Nag.) pseudo-R^2^, and root mean squared error (RMSE) remain unchanged—it only rescales the magnitude of the coefficients. All statistical analyses were performed in R (version 4.4.2); Negative binomial models were fitted using the MASS package. All maps were produced using ArcGIS Pro (version 3.4.0, Esri).

## 3. Results

### 3.1. Spatial and Temporal Patterns of Coccidioidomycosis Incidence and Climate Drivers

From 2000 to 2022, coccidioidomycosis incidence displayed noticeably spatial variability across Arizona, with highest incidence concentrated in Arizona’s southcentral counties—Maricopa, Pima, and Pinal (Figure 1a). Monthly incidence in these counties (Figure 1b) showed pronounced seasonal fluctuations and an overall long-term increasing trend, even after adjusting for artificial case inflation (Jun 2009–Nov 2012) and reporting delays. Despite these adjustments, each county in the study region still reported over 9,890 cumulative cases and a mean annual incidence exceeding 90 cases per 100,000 population, together accounting for ∼95% of statewide cases (135,210 out of 142,533).

Mean annual maps of climatic drivers from 1997 to 2022 showed distinct geographic variation that closely aligned with observed incidence patterns (Figure 3). The hyperendemic tri-county region was characterized by drier and hotter surface soil conditions (Figures 3a–3b) and the highest annual counts of dusty days (Figure 3c). Interestingly, Yuma County had the driest and hottest soil statewide but reported the lowest incidence. Wind patterns showed an unexpected inverse trend: the two highest-incidence counties—Maricopa (114 per 100,000) and Pinal (116 per 100,000)—had the lowest mean annual wind speeds (Figure 3d). In contrast, northeastern high-elevation counties with cooler, wetter soils, fewer dusty days, and stronger winds reported substantially lower incidence rates.

The mean monthly patterns of coccidioidomycosis incidence and climate variables (Figure 2) informed the seasonal groupings defined in the Methods. Incidence displayed a bimodal distribution, with a primary peak in November, a secondary peak in June–July, and a minimum in August followed by a secondary low in February–March. Largely, monthly dusty day counts mirrored incidence trends and were inversely related to topsoil moisture.

### 3.2. Multivariable Analysis of Climate Drivers on Coccidioidomycosis Incidence

Table 2 presents concurrent and lagged associations between seasonal coccidioidomycosis incidence and climate drivers, reported as aIRRs with 95% CIs and standardized coefficients (β), derived from MNBR inferential models. The table also shows predictors selected in the seasonal predictive models, which share identical aIRRs, β, and *p*-values with those from the corresponding inferential models. Most predictors retained in the predictive models were statistically significant (*p* < 0.1), and the majority of significant exposures identified in the inferential models were also selected in the prediction models. We restrict interpretation to statistically significant associations, excluding non-significant results (p ≥ 0.1 with CIs spanning 1), which imply no meaningful effect and help avoid spurious conclusions. For clarity, we use the terms “incidence season,” “seasonal incidence,” “exposure season,” and “modeling season” interchangeably throughout this study to refer to the season during which pathogen exposure occurs. Lag 0–12 refers to the SM or ST season occurring from the concurrent season (lag 0) up to 12 lagged seasons prior to the exposure season. In our lagged season-year notation, for example, “Winter-Y2 SM” refers to surface soil moisture during the winter season 2 years prior to the exposure season, while “Winter-Y0 SM” refers to winter soil moisture in the same year as the exposure season. The same notation applies to soil temperature in the topsoil.

**Table 2.** Multivariable Analysis of Coccidioidomycosis Incidence in Relation to Climate Drivers.

| Measure | Fall |  |  | Monsoon |  |  | Spring |  |  | Winter |  |  |
| --- | --- | --- | --- | --- | --- | --- | --- | --- | --- | --- | --- | --- |
| | aIRR<sup>a</sup> | $\beta^{e,f}$ | Lag<sup>g</sup> | aIRR<sup>a</sup> | $\beta^{e,f}$ | Lag<sup>g</sup> | aIRR<sup>a</sup> | $\beta^{e,f}$ | Lag<sup>g</sup> | aIRR<sup>a</sup> | $\beta^{e,f}$ | Lag<sup>g</sup> |
| Dusty days | 1.005 (1.0004, 1.01)<sup>c</sup> | **0.12** | 0 | 1.004 (1.001, 1.007)<sup>c</sup> | **0.10** | 0 | 1.005 (1.003, 1.008)<sup>c</sup> | **0.13** | 0 | 1.0004 (0.996, 1.004) | 0.01 | 0 |
| Wind speed | 1.30 (1.02, 1.64)<sup>c</sup> | **0.24** | 0 | 0.99 (0.85, 1.16) | -0.005 | 0 | 0.69 (0.53, 0.90)<sup>c</sup> | **-0.35** | 0 | 1.32 (1.06, 1.64)<sup>c</sup> | **0.26** | 0 |
| Soil moisture |  |  |  |  |  |  |  |  |  |  |  |  |
| Fall-Y0 | 0.93 (0.90, 0.96)<sup>b</sup> | **-0.26** | 0 |  |  | NA |  |  | NA |  |  | NA |
| Monsoon-Y0 | 1.03 (1.00, 1.07)<sup>d</sup> | **0.11** | 1 | 1.002 (0.97, 1.04) | 0.01 | 0 |  |  | NA |  |  | NA |
| Spring-Y0 | 0.88 (0.81, 0.95)<sup>c</sup> | **-0.47** | 2 | 0.91 (0.85, 0.97)<sup>c</sup> | **-0.34** | 1 | 0.92 (0.86, 0.98)<sup>c</sup> | **-0.30** | 0 |  |  | NA |
| Winter-Y0 | 1.10 (1.05, 1.15)<sup>b</sup> | **0.34** | 3 | 1.12 (1.08, 1.17)<sup>b</sup> | **0.41** | 2 | 1.04 (1.00, 1.08)<sup>d</sup> | **0.13** | 1 | 0.96 (0.92, 0.99)<sup>c</sup> | **-0.16** | 0 |
| Fall-Y1 | 1.01 (0.98, 1.04) | 0.03 | 4 | 0.95 (0.92, 0.98)<sup>c</sup> | **-0.20** | 3 | 0.94 (0.90, 0.97)<sup>c</sup> | **-0.23** | 2 | 0.98 (0.95, 1.01) | -0.07 | 1 |
| Monsoon-Y1 | 1.01 (0.98, 1.05) | 0.04 | 5 | 0.997 (0.96, 1.04) | -0.01 | 4 | 1.02 (0.98, 1.07) | 0.08 | 3 | 1.04 (1.01, 1.08)<sup>c</sup> | **0.15** | 2 |
| Spring-Y1 | 0.84 (0.77, 0.91)<sup>b</sup> | **-0.62** | 6 | 0.86 (0.80, 0.91)<sup>b</sup> | **-0.55** | 5 | 0.85 (0.80, 0.91)<sup>b</sup> | **-0.57** | 4 | 0.88 (0.82, 0.95)<sup>c</sup> | **-0.44** | 3 |
| Winter-Y1 | 1.11 (1.06, 1.16)<sup>b</sup> | **0.35** | 7 | 1.05 (1.02, 1.09)<sup>c</sup> | **0.19** | 6 | 1.12 (1.07, 1.18)<sup>b</sup> | **0.41** | 5 | 1.15 (1.10, 1.20)<sup>b</sup> | **0.48** | 4 |
| Fall-Y2 | 0.93 (0.90, 0.97)<sup>c</sup> | **-0.25** | 8 | 0.99 (0.96, 1.02) | -0.02 | 7 | 0.93 (0.90, 0.97)<sup>b</sup> | **-0.25** | 6 | 0.97 (0.93, 1.01) | **-0.11** | 5 |
| Monsoon-Y2 | 1.12 (1.07, 1.17)<sup>b</sup> | **0.39** | 9 | 1.02 (0.97, 1.06) | 0.06 | 8 | 0.99 (0.93, 1.04) | -0.05 | 7 | 1.09 (1.04, 1.15)<sup>c</sup> | **0.31** | 6 |
| Spring-Y2 | 0.91 (0.84, 0.98)<sup>c</sup> | **-0.34** | 10 | 0.82 (0.77, 0.88)<sup>b</sup> | **-0.69** | 9 | 0.77 (0.72, 0.82)<sup>b</sup> | **-0.93** | 8 | 0.79 (0.74, 0.85)<sup>b</sup> | **-0.80** | 7 |
| Winter-Y2 | 1.12 (1.08, 1.17)<sup>b</sup> | **0.40** | 11 | 1.11 (1.07, 1.14)<sup>b</sup> | **0.35** | 10 | 1.12 (1.08, 1.16)<sup>b</sup> | **0.39** | 9 | 1.16 (1.11, 1.20)<sup>b</sup> | **0.52** | 8 |
| Fall-Y3 | 0.94 (0.90, 0.98)<sup>c</sup> | **-0.21** | 12 | 0.96 (0.93, 0.99)<sup>c</sup> | **-0.16** | 11 | 0.95 (0.92, 0.98)<sup>c</sup> | **-0.18** | 10 | 0.97 (0.93, 1.01)<sup>d</sup> | **-0.12** | 9 |
| Monsoon-Y3 |  |  | NA | 1.10 (1.05, 1.15)<sup>b</sup> | **0.32** | 12 | 1.09 (1.05, 1.14)<sup>b</sup> | **0.31** | 11 | 1.08 (1.02, 1.14)<sup>c</sup> | **0.27** | 10 |
| Spring-Y3 |  |  | NA |  |  | NA | 0.99 (0.94, 1.05) | -0.02 | 12 | 0.93 (0.87, 0.99)<sup>c</sup> | **-0.26** | 11 |
| Winter-Y3 |  |  | NA |  |  | NA |  |  | NA | 1.07 (1.03, 1.11)<sup>c</sup> | **0.22** | 12 |
| Soil temperature |  |  |  |  |  |  |  |  |  |  |  |  |
| Fall-Y0 | 1.03 (0.94, 1.13) | 0.24 | 0 |  |  | NA |  |  | NA |  |  | NA |
| Monsoon-Y0 | 0.93 (0.84, 1.02) | -0.59 | 1 | 0.98 (0.91, 1.05) | -0.16 | 0 |  |  | NA |  |  | NA |
| Spring-Y0 | 1.11 (1.001, 1.24)<sup>c</sup> | **0.86** | 2 | 1.15 (1.05, 1.25)<sup>c</sup> | **1.09** | 1 | 1.11 (0.993, 1.23)<sup>d</sup> | **0.81** | 0 |  |  | NA |
| Winter-Y0 | 0.87 (0.81, 0.94)<sup>c</sup> | **-1.07** | 3 | 0.98 (0.92, 1.04) | -0.18 | 2 | 0.97 (0.90, 1.04) | -0.26 | 1 | 0.91 (0.84, 0.98)<sup>c</sup> | **-0.79** | 0 |
| Fall-Y1 | 1.27 (1.15, 1.40)<sup>b</sup> | **1.90** | 4 | 1.16 (1.05, 1.28)<sup>c</sup> | 1.18 | 3 | 1.16 (1.06, 1.26)<sup>c</sup> | **1.15** | 2 | 1.16 (1.03, 1.30)<sup>c</sup> | **1.18** | 1 |
| Monsoon-Y1 | 0.98 (0.90, 1.06) | -0.16 | 5 | 1.06 (0.96, 1.18) | 0.46 | 4 | 1.05 (0.94, 1.17) | 0.37 | 3 | 1.21 (1.11, 1.32)<sup>b</sup> | 1.50 | 2 |
| Spring-Y1 | 1.13 (1.03, 1.23)<sup>c</sup> | **0.95** | 6 | 0.98 (0.89, 1.07) | -0.20 | 5 | 0.996 (0.91, 1.09) | -0.03 | 4 | 1.18 (1.09, 1.29)<sup>c</sup> | **1.34** | 3 |
| Winter-Y1 | 1.15 (1.04, 1.27)<sup>c</sup> | **1.09** | 7 | 1.10 (1.03, 1.18)<sup>c</sup> | **0.74** | 6 | 0.99 (0.90, 1.09) | -0.07 | 5 | 0.86 (0.79, 0.93)<sup>c</sup> | **-1.19** | 4 |
| Fall-Y2 | 0.93 (0.85, 1.01) | -0.57 | 8 | 0.99 (0.91, 1.07) | -0.09 | 7 | 0.98 (0.90, 1.08) | -0.13 | 6 | 1.06 (0.96, 1.18) | 0.48 | 5 |
| Monsoon-Y2 | 0.96 (0.87, 1.06) | -0.32 | 9 | 0.97 (0.89, 1.07) | -0.20 | 8 | 0.96 (0.85, 1.09) | -0.33 | 7 | 1.03 (0.90, 1.16) | 0.20 | 6 |
| Spring-Y2 | 0.998 (0.90, 1.11) | -0.01 | 10 | 1.14 (1.07, 1.21)<sup>b</sup> | **1.03** | 9 | 1.01 (0.93, 1.10) | 0.11 | 8 | 1.097 (1.001, 1.202)<sup>c</sup> | 0.74 | 7 |
| Winter-Y2 | 0.91 (0.83, 1.002)<sup>d</sup> | -0.76 | 11 | 1.05 (0.96, 1.15) | 0.37 | 10 | 1.10 (1.03, 1.19)<sup>c</sup> | **0.79** | 9 | 1.16 (1.06, 1.27)<sup>c</sup> | **1.18** | 8 |
| Fall-Y3 | 0.92 (0.82, 1.03) | **-0.68** | 12 | 1.01 (0.93, 1.11) | 0.11 | 11 | 0.999 (0.91, 1.10) | -0.01 | 10 | 0.99 (0.90, 1.08) | -0.11 | 9 |
| Monsoon-Y3 |  |  | NA | 0.89 (0.83, 0.96)<sup>c</sup> | **-0.88** | 12 | 0.84 (0.76, 0.93)<sup>c</sup> | **-1.39** | 11 | 1.18 (1.06, 1.32)<sup>c</sup> | **1.33** | 10 |
| Spring-Y3 |  |  | NA |  |  | NA | 1.003 (0.93, 1.08) | 0.02 | 12 | 1.06 (0.98, 1.15) | **0.46** | 11 |
| Winter-Y3 |  |  | NA |  |  | NA |  |  | NA | 1.07 (0.99, 1.16)<sup>d</sup> | 0.55 | 12 |

*Note.* Fall-Y0 refers to the fall soil moisture (SM) or soil temperature (ST) season occurring in the same year as the incidence season; Winter-Y3 refers to the winter SM/ST occurring 3 years prior to the incidence season, and so forth, with seasons ordered from most to least recent. N/A (not applicable) indicates seasons that fall outside the analysis window (i.e., before the concurrent season or beyond 3 years prior). <sup>a</sup>Multivariable negative binomial regression (MNBR) inferential models with the Keeping-Exposure Stepwise Selection (KESS) approach were used to estimate adjusted incidence rate ratios (aIRRs) along with their 95% confidence intervals (95% CIs). <sup>b</sup> $p$ -value < 0.0001. <sup>c</sup>0.0001 ≤ $p$ -value < 0.05. <sup>d</sup>0.05 ≤ $p$ -value < 0.1. <sup>e</sup>Standardized coefficients ( $\beta$ ) from the KESS-selected MNBR inferential models were reported, using z-score-standardized predictors. <sup>f</sup>Bolded $\beta$ coefficients denote predictors retained in the seasonal predictive models, which were selected via an unrestricted stepwise MNBR procedure minimizing AIC. Because the predictive and inferential models share the same stepwise selection and covariates set, these retained predictors have identical aIRRs, $\beta$ , and $p$ -values in both model types.
<sup>g</sup>Seasonal lags are labeled sequentially from concurrent (0) to 12 seasons prior.

Across seasons, SM displayed significant associations with incidence in more concurrent and lagged periods than ST; however, in the seasonal predictive models, individual ST exposures generally showed greater influence on incidence predictions compared to individual SM exposures. SM–incidence associations followed an alternating wet–dry pattern, with drier surface soils in the concurrent season for fall, spring, and winter, and one season prior to monsoon exposure, linked to increased incidence. Comparatively, ST–incidence associations generally followed a cool–hot cycle, with significant effects emerging at lag 0 for spring and winter incidence, lag 1 for monsoon, and lag 2 for fall.

Significant associations between incidence and both concurrent and lagged SM were largely consistent across seasonal models and SM seasons. Specifically, increased incidence was associated with wetter surface soils during monsoon and winter seasons, and with drier conditions in fall and spring seasons occurring 0–3 years prior to the exposure season. Although lagged wetter winters were associated with increased incidence, concurrent wetter seasons— including the concurrent winter—were associated with decreased incidence. Comparatively, associations between ST and incidence showed greater variability among seasonal models and ST seasons. Elevated ST conditions in spring seasons 0–2 years prior to the exposure season, as well as hotter fall soils one year prior, were associated with increased incidence. Monsoon ST effects varied by exposure timing: higher ST one and three years prior elevated winter incidence, while lower ST three years prior elevated spring and monsoon incidence. Winter ST showed mixed associations with incidence: cooler soils in the exposure year and hotter soils 3 years prior increased incidence, one-year-prior ST was positively associated with fall and monsoon incidence but negatively with winter incidence, and two-year-prior ST was negatively linked to fall yet positively to spring and winter incidence. Wind speed was positively associated with incidence in fall and winter, but negatively in spring. The number of dusty days was positively associated with incidence in fall, monsoon, and spring, and showed a positive but non-significant association in winter.

Spring and winter soil conditions—both SM and ST—contributed most significantly to coccidioidomycosis incidence. Notably, all winter and nearly all spring SM exposures from 0 to 3 years prior were significantly associated with incidence across every seasonal model (with only Spring-Y3 for spring incidence as an exception) and showed larger absolute coefficients (|β|).

Winter incidence was particularly sensitive to soil conditions, with more significant SM and ST associations than any other season. All winter SM and ST exposures, as well as all monsoon and spring SM exposures, were significantly associated with winter incidence, whereas fall soil conditions had the least influence. Overall, increased winter incidence was linked to drier and hotter fall and spring conditions, wetter and hotter monsoons, drier and cooler concurrent winter, wetter and cooler winter one year prior, and wetter and hotter winters 2 to 3 years prior.

Spring incidence was less sensitive to ST conditions than any other season, with only 4 significant ST lags. Comparatively, ten SM lags were significantly associated with spring incidence, with spring SM—particularly two years prior (β, −0.93; aIRR, 0.77 [95% CI, 0.72-0.82])—along with three-year-prior monsoon ST and one-year-prior fall ST, showing the strongest influence on spring incidence prediction. Overall, increased spring incidence was associated with drier and hotter fall and spring seasons, wetter and cooler monsoon 3 years prior, wetter winters 0 to 2 years prior—particularly a warmer winter two years prior.

Monsoon incidence was driven less by monsoon SM conditions than to SM in other seasons. Nine SM lags and five ST lags showed significant associations, with spring and winter SM, one-year-prior fall ST, and spring ST at 0 and 2 years prior showing the strongest predictive influence. Overall, higher monsoon incidence followed drier and hotter falls and springs, wetter and cooler monsoon 3 years prior, and wetter winters—particularly a warmer winter one year prior.

Fall incidence was associated with 11 SM and 6 ST lags, primarily from non-monsoon seasons. Monsoon ST showed no significant effects, and only monsoon SM at 0 and 2 years prior showed positive associations. In contrast, all winter SM and ST lags and all spring SM lags were significantly associated with fall incidence. The most influential predictor was one-year-prior fall ST (β, 1.90; aIRR, 1.27 [95% CI, 1.15-1.40]). Overall, elevated fall incidence was linked to drier and hotter falls and springs, wetter monsoons at 0 and 2 years prior, wetter and cooler winters at 0 and 2 years prior, and wetter and warmer winter one year prior.

### 3.3. Evaluation of Seasonal Prediction Models

Table 3 shows that all four seasonal models explain a significantly high proportion of incidence variance in the full-data fit (adj. R² = 0.94–0.95; Nag. pseudo-R² = 0.79–0.83) and maintain similar predictive performance under LOOCV (adj. R² = 0.88–0.90; Nag. R² = 0.79–0.83). The minimal discrepancy between in-sample and cross-validated metrics indicates negligible overfitting and strong generalizability. Predictive performance was consistently strong across all seasons, with little variation among the models.

**Table 3.** Performance Metrics for Seasonal Prediction Models.

| Season | In-sample model (full-data fit) |  |  |  | Cross-validated model (LOOCV) |  |  |  |
| --- | --- | --- | --- | --- | --- | --- | --- | --- |
| | AIC | $R^2$ | Adj. $R^2$ | Nag. $R^2$ | $R^2$ | Adj. $R^2$ | Nag. $R^2$ | RMSE |
| Fall | 806.80 | 0.96 | 0.95 | 0.81 | 0.92 | 0.89 | 0.82 | 3.77 |
| Monsoon | 768.81 | 0.95 | 0.94 | 0.79 | 0.91 | 0.89 | 0.79 | 3.00 |
| Spring | 772.83 | 0.96 | 0.95 | 0.81 | 0.93 | 0.90 | 0.81 | 2.70 |
| Winter | 763.81 | 0.96 | 0.95 | 0.83 | 0.91 | 0.88 | 0.83 | 2.90 |

*Note.* Seasonal stepwise MNBR predictive models were evaluated using (1) in-sample full-data fit—reporting Akaike Information Criterion (AIC), coefficient of determination ( $R^2$ ), adjusted $R^2$ , and Nagelkerke (Nag.) pseudo- $R^2$ —and (2) out-of-sample performance via leave-one-out cross-validation (LOOCV), reporting $R^2$ , adj. $R^2$ , Nag. $R^2$ , and root mean squared error (RMSE). All seasonal models achieved statistical significance ( $p$ -value < 0.0001).

Figures 4a–4l present the time series of observed and LOOCV-predicted coccidioidomycosis incidence for each season and county within the study region from 2000 to 2022. In all cases, the models closely reproduce the temporal patterns—capturing major peaks and troughs in incidence—demonstrating strong agreement between predicted and observed trends. The primary exception was the monsoon model for Pinal County from 2003 to 2005, which, although approximating the magnitude of incidence, failed to capture the slight declining trend and instead predicted an upward trend.

**Figure 4.**
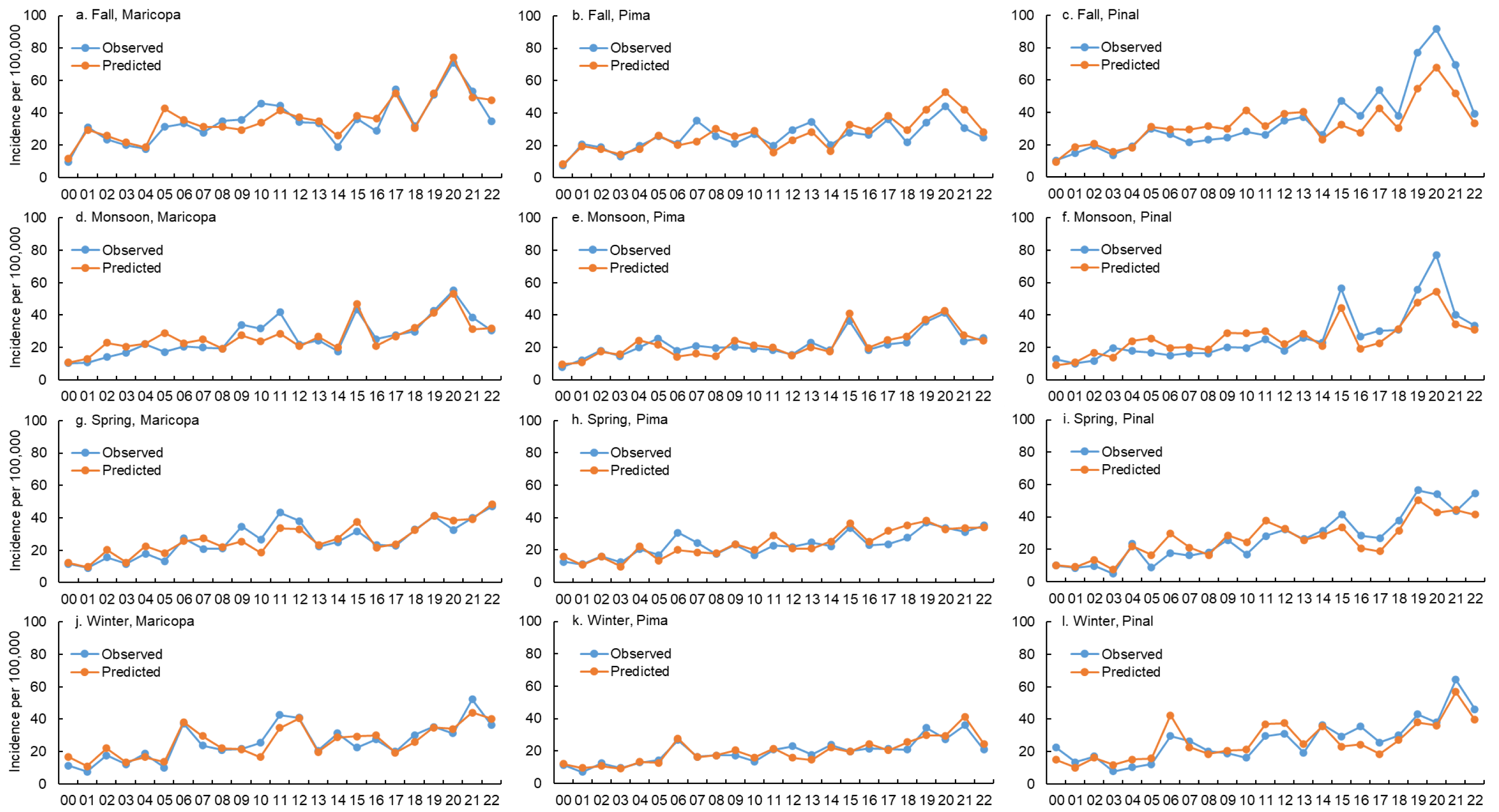
Observed versus predicted seasonal coccidioidomycosis incidence in Maricopa, Pima, and Pinal counties, 2000–2022. Predictions were generated via LOOCV of unrestricted stepwise MNBR predictive models across the four defined seasons: Fall (a–c), Monsoon (d–f), Spring (g–i), and Winter (j–l). Observed incidence is shown in blue (lines and circles); predicted incidence in orange.

## 4. Discussion

### 4.1. Spatial Patterns—Climate Niches of Coccidioidomycosis in Arizona

Annual maps showed distinct climate niches associated with coccidioidomycosis incidence across Arizona. The highest incidence rates were concentrated in Maricopa, Pima, and Pinal counties, which exhibited drier, hotter surface soils and more frequent dusty days. These findings align with prior research linking coccidioidomycosis hot spots to the low-elevation Sonoran Desert’s hot, arid, and dust-prone environment (Gorris et al., 2018). Yet Yuma County—despite having the driest and hottest soils in the state—recorded the lowest incidence, suggesting that excessively arid and hot environments may inhibit *Coccidioides* growth and that the fungus thrives within a specific climatic threshold. Although *Coccidioides* can survive across a broad temperature range under laboratory conditions (Friedman et al., 1956), optimal growth is believed to occur between 20°C and 40°C, with growth declining sharply at higher soil temperatures—particularly under very dry conditions (Fisher et al., 2007). Conversely, the northeastern high-elevation counties—characterized by cooler, wetter soils, fewer dusty days, and stronger mean winds—exhibited the lowest incidence rates, implying that mean wind alone is insufficient to drive exposure risk. Together, these findings highlight the complexity of climatic influences shaping coccidioidomycosis endemicity in Arizona.

### 4.2. Role of Climate Drivers in Coccidioidomycosis Dynamics

#### 4.2.1. Consistency of Soil Moisture Versus Soil Temperature Effects Across Years and Exposure Seasons

Multivariable regression results indicated that soil moisture was a more widespread and consistent driver of coccidioidomycosis incidence than soil temperature across seasons and years. In all four exposure seasons, SM showed significant associations with incidence in roughly twice as many time periods as ST. This finding aligns with a previous study in Kern County, California (CA), by Weaver and Kolivras (2018), which used precipitation and air temperature (AT) as proxies for SM and ST, respectively, and found precipitation to be the more frequently significant predictor—about 3.5 times more often than AT. However, our models explained a much larger proportion of incidence variance and demonstrated stronger generalizability, likely reflecting the improved climate representation achieved with direct SM and ST indicators over proxy measures.

Across all four incidence seasons, concurrent and lagged SM showed consistent directional effects within each season across the years (e.g., spring SM from 0 to 3 years prior was consistently negatively associated with incidence across exposure seasons), while ST effects were more variable in Monsoon and Winter lags. In general, SM and ST during spring and winter were more influential on incidence and showed more significant lags than in the other seasons. Specifically, wetter soils in winters and monsoons, and drier, hotter soils in falls and springs, were associated with higher incidence. These findings support the “grow and blow” hypothesis by implying that adequate soil moisture in winter and monsoon seasons promotes hyphal growth, while heat and dryness in fall and spring facilitate hyphal desiccation, arthroconidia formation, and dispersal. Weaver and Kolivras (2018) similarly found that winter and spring precipitation were the most commonly significant predictors and contributed most to increased incidence. A recent study of 14 hyperendemic counties in California (Head et al., 2022) also found that within a transmission season, higher fall incidence was associated with higher precipitation and lower AT in winter and spring, but with lower precipitation and higher AT in summer and fall. Within a year, our results showed that higher fall incidence was associated with higher SM and lower ST in winter and monsoon, but with lower SM and higher ST in spring and fall. Actually, our findings for SM align with the two CA studies, with the explanation that wetter conditions during typically dry seasons may inhibit dust production and thereby reduce spore dispersal (summer and fall are typically dry in CA versus spring and fall in our study region).

#### 4.2.2. Seasonal Incidence Differentially Shaped by Soil Moisture and Soil Temperature

Both SM and ST showed significant influence on incidence across all exposure seasons for up to three years prior—a pattern consistent with previous studies, including those in CA using precipitation and AT (Head et al., 2022), and in Pima County, AZ using precipitation (Comrie, 2005) or NDVI (Stacy et al., 2012) as proxies. While we also used linear modeling like the two AZ studies, our models identified many more significant associations with stronger performance, which may be due to direct SM data instead of proxies, a larger sample size, and the inclusion of both soil hydrothermal drivers (SM and ST). Despite consistent SM effects within each season across years and similar ST effects in fall and spring lags, the influence of soil moisture and soil temperature on incidence varies discernibly across incidence seasons. Winter incidence was most sensitive to soil hydrothermal conditions, with 20 significant lags (11 SM, 9 ST), followed by fall (11 SM, 6 ST), spring (10 SM, 4 ST), and monsoon (9 SM, 5 ST). This finding aligns with Stacy et al. (2012), who found January–April incidence most sensitive to NDVI lags, and with Weaver and Kolivras (2018), who reported winter and fall incidence most responsive to precipitation and AT. However, unlike Weaver and Kolivras (2018) and Comrie (2005), our best-performing models did not align with peak-incidence seasons, as also shown by Stacy et al. (2012). Instead, winter (lowest incidence) and fall (highest) models showed similarly strong explanatory power—likely due to differences in hydrothermal metrics used.

#### 4.2.3. Concurrent Dispersion Conditions

Our results indicate that concurrent dispersion conditions played an important but seasonally distinct role: fall (lower SM, more dusty days, higher wind speed), monsoon (more dusty days), spring (lower SM, higher ST, more dusty days, lower wind speed), and winter (lower SM, lower ST, higher wind speed). The consistent positive association between number of dusty days and incidence across exposure seasons—and the alignment of its temporal trends and spatial distribution with monthly and annual incidence patterns (Figures 1–3; Table 2)—support the idea that more dust increases exposure risk. This finding supports earlier AZ studies—Comrie (2005) found positive PM_10_–incidence associations in foresummer (May–Jul) and winter (Jan–Apr), and Tong et al. (2017) observed positive correlation between dust frequency and incidence—and recent research by Weaver et al. (2025), which identified a positive exposure–response relationship between cumulative fine mineral dust exposure 1–3 months prior to disease onset and incidence across 14 hyperendemic CA counties. The Southwest is projected to experience increased dust emissions alongside greater aridity due to human-caused climate change (Achakulwisut et al., 2019), which is expected to elevate coccidioidomycosis risk. We found a positive association between wind speed and incidence in fall and winter, but a negative one in spring—an unexpected inverse pattern also appeared in annual wind and incidence maps.

Similarly, Weaver and Kolivras (2018) reported that concurrent mean 5-s gusts correlated positively with winter incidence, while concurrent 2-min gusts correlated negatively with summer incidence. In arid, dust-prone spring, dispersion seems driven mainly by dry, hot soils and high dustiness; mean seasonal wind alone may not suffice, as exposure risk can remain high even under low mean seasonal wind, given increased outdoor activities or strong, brief gusts.

Future work should explore additional soil-disturbance metrics, such as the frequency of hourly peak wind gusts exceeding erosion thresholds at fine spatial scales, wildfire occurrence, patterns of outdoor nature-based recreation (e.g., desert hiking, backcountry use, and park visitation), and agricultural and construction activities.

Across exposure seasons, lower concurrent SM was consistently linked to higher incidence, conforming to Porter et al. (2024), who reported a negative association between mean daily soil moisture and airborne *Coccidioides* detection—suggesting that dryness promotes spore aerosolization. Concurrent ST was positively linked to incidence in the dry spring but negatively in the wet winter. This unexpected dry–cool pattern in typically wet, low-UV (EPA, 2025; Lin & and Li, 2002) winters may reflect: (a) dryness and high winds were likely the primary concurrent dispersion drivers, (b) seasonal models may not have captured differing ST effects between early and late winter, or (c) lower temperatures are linked to reduced UV exposure (Kadad et al., 2020), which may prolong spore survival in the air or on surfaces. Friedman et al. (1956) found greater *Coccidioides* viability at –15°C than 4°C after six months under high humidity, but they only tested limited temperatures, whereas Arizona tri-county winter temperatures averaged 8– 20 °C (NOAA, 2025). Further laboratory studies across finer temperature–humidity gradients are therefore needed.

#### 4.2.4. Soil Hydrothermal Cycles in the Context of Prevailing Hypotheses

Collectively, although no single “recipe” of climate conditions explains all seasonal incidence patterns, our findings support that alternating wet–dry and cool–hot cycles along with concurrent dry, dusty conditions may significantly influence coccidioidomycosis incidence. Our results align with Head et al. (2022), who found that fall incidence was linked to prior wet–dry (via precipitation) and cool–hot (via AT) cycles over a three-year period. We generalized this pattern by demonstrating similar hydrothermal cycles across all incidence seasons and by incorporating concurrent dispersion factors (e.g., dry, windy, and dusty conditions during fall). Unlike previous precipitation-based analyses in AZ (Comrie, 2005) and CA (Head et al., 2022), our models using Noah-simulated SM showed more consistent associations across years and exposure seasons.

This likely reflects the soil’s buffering capacity—where infiltration, drainage, and evapotranspiration smooth out rainfall pulses—resulting in lower variability and greater persistence of SM compared to precipitation (Ghannam et al., 2016). Interestingly, increased incidence was significantly associated with prior dry–hot, wet–hot, and wet–cool surface soil conditions, but not with dry–cool conditions. This suggests that moisture or heat—at different points in the prior multi-year cycle—was necessary to support fungal growth or dispersal, while a sustained absence of both did not appear beneficial to the fungus.

Three prevailing hypotheses may complementarily explain these multi-year soil hydrothermal cycles: “grow and blow”, “soil sterilization”, and “endozoan, small-mammal reservoir” hypotheses (Comrie, 2005; Fisher et al., 2007; Maddy, 1965; Taylor & Barker, 2019). The temporal sequence suggested by our findings is: (a) sufficient soil moisture during prior winter and monsoon seasons likely promoted hyphal growth (“grow”) and increased plants that sustained small mammal populations (“endozoan”); (b) subsequent hot–dry fall and spring seasons might suppress other soil microbes competing with *Coccidioides* (“sterilization”), but enhance arthroconidial fragmentation and dispersal (“blow”), potentially infecting more naive animals (“endozoan”); (c) when favorable hydrothermal conditions return subsequently, *Coccidioides* hyphae might thrive with reduced microbial competition (“sterilization”), and endospores released from infected dead hosts could covert to hyphae using carcasses as a nutrient source (“endozoan”); (d) Later on, favorable concurrent dispersion conditions—dry, windy, and dusty—may finally increase human infection risk (“blow”).

### 4.3. Limitations

This study has several limitations. First, coccidioidomycosis case counts were subject to exposure misclassification bias due to: (a) cases being counted by area of residence rather than the actual exposure location, (b) delays between exposure date and report date. To reduce bias from (a)—cases acquired outside the residential county due to work or travel—we limited analyses to the hyperendemic tri-county region. To limit (b), we applied a 1-month offset to adjust for reporting delays and conducted analyses at a broad temporal scale (season). Second, case counts may be biased by changes in reporting and diagnostic practices, and public or clinical awareness. To correct for abnormal case inflation from 2009 to 2012—due to changes in testing and reporting—we halved the counts during that period. Third, the reanalysis-derived SM and ST data carried uncertainties from model parameterizations, may not capture micro-scale soil heterogeneity, and can differ from direct in-situ measurements. However, our analyses were conducted at a broad spatial scale (county), and NLDAS-2 SM and ST data have been rigorously validated and demonstrated to effectively capture and align well with patterns of observed values (Xia et al., 2013, 2014). Fourth, seasonal analyses cannot distinguish the effects of exposure occurring at different times within a season. Fifth, our longitudinal ecological design precludes causal inference, and is subject to ecological fallacy, which can lead to incorrect inferences about individuals.

### 4.4. Future Directions

Since *C. immitis* dominates in CA and Washington state, while *C. posadasii* is prevalent in AZ and other endemic regions—with little spatial overlap between the two species (Ashraf et al., 2020; Head et al., 2022)—it is important to assess whether the SM and ST relationships we identified also apply in CA. Due to limited sample size, we did not use more complex models; future research should explore nonlinear methods to estimate exposure–response relationships and compare them with our findings. We identified significant multi-year climate–incidence associations, suggesting that prior long-term soil hydrothermal trends could influence *Coccidioides* dynamics. Climate projections indicate continued warming, intensified drought, and more frequent extreme rainfall events (Cook et al., 2015, 2021; Diffenbaugh & Giorgi, 2012; A. P. Williams et al., 2020, 2022), all of which would potentially alter soil hydrothermal cycles. Applying these projections to evaluate the potential expansion of endemic regions would therefore be valuable. In addition, future studies should examine how SM and ST influence the timing, intensity, and duration of coccidioidomycosis transmission seasons, which might shift in response to evolving climate conditions. Finally, establishing a long-term surveillance network for *Coccidioides*—along with meteorological, soil, and human and animal activity data—would significantly advance our understanding of fungal ecology and disease dynamics. Such a system is essential for improving coccidioidomycosis forecasting and preparing endemic communities for the challenges of a warming, drying, and increasingly variable climate.

## 5. Conclusions

To our knowledge, this is the first study to comprehensively evaluate the concurrent and lagged effects of soil moisture and soil temperature on coccidioidomycosis incidence. We found that incidence was higher in areas with drier, hotter surface soils and more frequent dusty days. Our findings underscore the critical role of multi-year soil hydrothermal cycles in shaping coccidioidomycosis dynamics. Multi-year alternating wet–dry and cool–hot cycles along with concurrent dry, dusty conditions, are associated with higher incidence across exposure seasons and support the prevailing “grow and blow,” “soil sterilization,” and “animal reservoir” hypotheses. Notably, we found no evidence linking prior dry–cool conditions to increased incidence, suggesting moisture or heat is essential prerequisite for the proliferation or dissemination of *Coccidioides*. Across exposure seasons, SM showed more widespread and stable seasonal effects than ST, with winter and spring soil conditions influencing the multi-year signal most heavily. Within annual cycles, increased incidence typically followed wetter winters and monsoons as well as drier, hotter springs and falls. Our models using NLDAS-2 SM and ST data effectively captured important seasonal and interannual variations in disease incidence, showing strong predictive performance and generalizability across exposure seasons. Our findings call for incorporating multi-year soil hydrothermal indicators—with up to a three-year lead time—into early-warning systems to enhance public health preparedness and response in endemic regions facing a growing burden of this climate-sensitive disease.

## Acknowledgments

We gratefully acknowledge Thomas Williamson and Guillermo Adame of the Arizona Department of Health Services for their assistance in obtaining coccidioidomycosis case data. We also thank JoEllyn McMillan, Cheng Zheng, Hongying Dai and Abraham Mengist (University of Nebraska Medical Center) and Azar Abadi (University of Alabama at Birmingham) for their valuable suggestions or comments on the research. This work was supported in part by the National Oceanographic and Atmospheric Administration’s National Integrated Drought Information System (Grant No. NA20 OAR4310368) and the National Aeronautics and Space Administration (NASA) Research Opportunities in Space and Earth Sciences (ROSES) program (Grant no. 8ONSSC22K1050).

## Conflict of Interest

The authors declare no conflicts of interest relevant to this study.

## Open Research

### Data Availability Statement

Coccidioidomycosis case-count data were provided by the Arizona Department of Health Services (ADHS) and can be obtained upon reasonable request from ADHS, subject to ADHS data-sharing policies. All other data used in this study are publicly available online, and their sources are listed in Table 1 (EPA, 2024; NLDAS project, 2022a, 2022b; Xia et al., 2012).

### Contributor Roles

Qianqian Li conducted conceptualization, disease and dust data curation, methodology development, formal analysis, project administration, validation, visualization, and original draft writing. Beichen Zhang contributed to the curation of NLDAS-2 climate data. Runqiu Wang, Haiyue Li, and Yue Zhan assisted with statistical methods. Daniel Tong assisted with methods for dust characterization. Jesse E. Bell acquired funding, supervised the study, and approved the final submission for publication. All authors reviewed and edited the final manuscript.

## Notes

### Competing Interest Statement

The authors have declared no competing interest.

### Author Declarations

Ethics committee/IRB of the University of Nebraska Medical Center (UNMC) waived ethical approval for this work. The work relied exclusively on de-identified, county-level coccidioidomycosis case-count data provided by the Arizona Department of Health Services (ADHS), which are available upon request from the ADHS. These case-count data contain no individual identifiable information or protected health information. And we did not interact or intervene with individuals to obtain information about them. Accordingly, the UNMC IRB Office of Regulatory Affairs (ORA) has determined that this project does not constitute human subject research as defined at 45CFR46.102 and is therefore not subject to the federal regulations.

